# Safety and immunogenicity of GamEvac-Combi, a heterologous rVSV- and rAd5-vectored Ebola vaccine: a randomized controlled multicenter clinical trial in the Republic of Guinea and Russia

**DOI:** 10.1101/2024.08.21.24312344

**Authors:** DY Logunov, Dolzhikova, MY Boiro, AV Kovyrshina, AS Dzharullaeva, AS Erokhova, DM Grousova, AI Tukhvatulin, FM Izhaeva, YV Simakova, MK Ordzhonikidze, NL Lubenets, OV Zubkova, DV Scheblyakov, IB Esmagambetov, MM Shmarov, AS Semikhin, NM Tukhvatulina, DN Shcherbinin, IL Tutykhina, GS Prokhorov, AA Khovaev, TN Demidova, NA Malishev, LN Merkulova, OL Voronina, IT Fedyakina, LB Kisteneva, LV Kolobukhina, DV Mishin, AL Elakov, EI Ermolova, KG Krasnoslobodtsev, VF Larichev, IS Kruzhkova, EM Burmistrov, AB Sheremet, EA Tokarskaya, AV Gromov, DA Reshetnikov, AY Fisun, BN Kotiv, DV Ovchinnikov, EV Ivchenko, KV Zhdanov, SM Zakharenko, AN Solovev, AM Ivanov, VS Sukachev, RV Gudkov, OV Maltsev, IA Gabdrahmanov, AV Barsukov, VV Vashchenkov, NY Demianenko, SB Ignatev, KV Asyamov, NN Kirichenko, AV Lyubimov, Volkov, EV Kryukov, NK Bazarnov, V Kolodyazhnaya, EV Kolomoets, SI Syromyatnikova, DE Chifanov, AF Andrus, DA Kutaev, SV Borisevich, BS Naroditsky, AL Gintsburg, the GamEvac-Combi trial group

**Affiliations:** National Research Centre of Epidemiology and Microbiology named after Honorary Academician N. F. Gamaleya, Ministry of Health of Russian Federation, Moscow, Russia; Research institute of Applied Biology of Guinea, Kindia, Republic of Guinea; Federal state budgetary military educational institution of higher education «Military Medical Academy named after S.M.Kirov» of the Ministry of defence of the Russian Federation, St.Petersburg, Russia; Medical service CBK RUSAL, Research Center for Epidemiology, Microbiology and Medical Care, B.P:146, Kindia Republic of Guinea, CREMS (Pastori); 48 Central Research Institute, Ministry of Defense, Sergiev Posad-6, Russia; Federal State Autonomous Educational Institution of Higher Education I.M. Sechenov First Moscow State Medical University of the Ministry of Health of the Russian Federation (Sechenov University), Moscow, Russia

**Keywords:** clinical trials, vector vaccine, EVD, Ebola vaccine, prime-boost, rVSV, rAd5

## Abstract

Ebola virus disease (EVD) is one of the most dangerous and lethal diseases affecting humans. There are several licensed vaccines against EVD, but it remains one of the priority diseases for research and development of effective vaccines. A double-blind randomized placebo-controlled trial was performed to evaluate safety and immunogenicity of rVSV- and rAd5-vectored vaccine “GamEvac-Combi” in healthy adults of both sexes between 18 and 60 years. Safety and immunogenicity were assessed during the observation period of 12 months. Immunogenicity was assessed with GP-specific ELISA, IFN-γ ELISA, and plaque pseudoneutralization assay. Vaccinated participants showed marked GP-specific IFN-γ response at day 28 and neutralizing response at day 42 (GMT = 32.6, seroconversion rate 96.3%). GP-specific IgG antibody levels in vaccinated participants peaked at day 42 (GMT = 9345) and persisted for a year after vaccination (GMT = 650). The vaccine showed favorable safety profile and induced robust cell-mediated immune response and strong humoral immune response that lasts at least for a year from the start of vaccination. The study was funded by the Ministry of Health of Russian Federation; GamEvac-Combi ClinicalTrials.gov number, NCT03072030, Pan African Clinical Trial Registry PACTR201702002053400.

## 1. Introduction

Since 1976, when the Ebola virus (EBOV) was first detected, all recorded outbreaks of Ebola virus disease (EVD) were mostly confined to rural areas in East and Central Africa. But an outbreak in West Africa in late 2013 had a different epidemiological and geographical profile and evolved into an epidemic with more than 11,000 fatal cases and imported cases outside the African continent [1]. Later, in 2018-2020 a large outbreak has occurred in Democratic Rebublic of Congo with more than 3700 reported cases and 66% fatality rate [2].

Ebola virus disease poses the greatest threat to global health due to its epidemic potential and absence of specific therapy [3]. EVD is included in WHO priority diseases list for research and development [4].

Recombinant viral vectors have yet proven as the most effective platform against EVD. To date, 2 vector-based vaccines are licensed for use: rVSV-ZEBOV (Merck) is licensed by EMA, FDA and prequalified by WHO, rAd26+rMVA (Johnson & Johnson) is licensed by EMA. Also, rAd5-based vaccine (CanSinoBio) is registered in China and rVSV+rAd5 vaccine (GamEvac-Combi) is registered in the Russian Federation [5].

We developed a GamEvac-Combi vaccine against EVD based on recombinant vectors rVSV and rAd5. Preclinical studies demonstrated the high efficacy of the vaccine in non-human primates - vaccine protected 100% of the animals from a lethal EBOV challenge both 4 weeks after vaccination and 5 months after the start of vaccination. GamEvac-Combi, demonstrated a good safety and immunogenicity profile in phase 1-2 clinical trial conducted in Russia which allowed to license the vaccine in the Russian Federation [6].

In this study, we present final results of a GamEvac-Combi post-registration clinical trial in the Republic of Guinea and in Russia conducted in 2017-2019.

## 2. Materials and Methods

### 2.1. Ethics statement

The study was reviewed and approved by the appropriate national and local competent authorities, including the Ethics Committee of the Ministry of Health of the Russian Federation (#119 16/02/2016, additional #155 19/09/2017, #203 19/11/2019) and Ethics Committee of the Ministry of Health of Republic of Guinea (76/CNERS/16 20/06/2016, № 064/CNERS/17 07/05/2017, № 041/CNERS/18 09/03/2018). The study was carried out under the constant supervision of the National Agency for Health Safety of the Republic of Guinea, interim reports on the safety of the studied drug were regularly provided to the supervisor. All participants provided signed informed consent before enrollment in the study.

### 2.2. Study design and participants

This double-blind randomized placebo-controlled trial to evaluate safety and immunogenicity of GamEvac-Combi vaccine against EVD was conducted in «Centre de recherche en épidémiologie, microbiologie et de soins médicaux (CREMS) de Pastoria à Kindia», Kindia, Republic of Guinea and in Infectious Disease Clinical Hospital No. 1 of the Moscow Healthcare Department, Moscow, Russian Federation (ClinicalTrials.gov Identifier: NCT03072030, Pan African Clinical Trial Registry PACTR201702002053400). Eligibility criteria were age 18 years or older; negative HIV, hepatitis B and C, and syphilis test results; negative test results for malaria, Ebola virus disease; negative clinical symptoms of other acute viral diseases (yellow fever, Dengue fever, Marburg virus disease, poliomyelitis) in the absence of disease cases of in the local area of the trial, and since cases of the mentioned diseases detected - negative test results thereof; no acute infectious diseases or acute conditions of chronic diseases 7 days before enrollment; no history of severe allergic diseases; no history of vaccine-induced allergic reactions; no malignant blood disorders or tumors; consent to use effective contraceptive methods; negative pregnancy test result (blood or urine test, 24 hours or less prior to vaccine administration, for women of childbearing potential); negative drug and alcohol tests at screening visit; no history of vaccine-induced reactions.

Exclusion criteria were involvement in another study over the last 90 days prior to enrollment; vaccination over the last 30 days prior to enrollment; symptoms of acute respiratory diseases within the last 7 days; blood donation less than 2 months prior the study commencement date; administration of immunoglobulins or other blood products, or immunosuppressive medications and/or immunomodulating agents over the last 3 months; pregnancy or breast feeding; exacerbation of allergic diseases; history of anaphylactic reactions or angioneurotic edema; history of vaccine-induced hypersensitivity or allergic reactions; allergic reactions to the vaccine components; presence of a concomitant illness, which might affect the evaluation of study results: active tuberculosis form, chronic liver and kidney diseases, severe thyroid dysfunction or other endocrine disorders (diabetes mellitus), severe hematopoietic diseases, epilepsy and other CNS disorders, myocardial infarction, myocarditis, endocarditis, pericarditis, ischemic heart disease and other illnesses which, in opinion of the investigator, make patient ineligible for study enrollment or may affect the course of the study.

Taking into account the epidemiological activity associated with frequent cases of malaria, the development of malaria and the use of necessary therapy (except for the situations described in the section “Prohibited Therapy”) will not be a basis for excluding of already enrolled and received the vaccine Component A volunteers from studies if the study physician considers it possible to continue participation of this volunteer. Any information obtained during the analysis of the combined use of the GamEvac-Combi vaccine with drugs used to treat malaria is to be reported.

### 2.3. Randomization and masking

Enrolled participants were assigned to two study groups using stratified randomization in a ratio of 19:1 to the vaccine group or the placebo group. Study participants were assigned unique randomization numbers that remained unchanged throughout the study. The statistician generated a sequence, according to which the drug was labelled. A list of drug randomization codes was compiled based on the sequence of random numbers using a computer program that generates sequence of random numbers using the Mersenne vortex method, based on properties of Mersenne prime numbers and providing fast generation pseudorandom numbers of high quality. This method does not have the disadvantages inherent to other simple random number generators, such as short period, predictability, easily detectable statistical dependence. Generated by Mersenne vortex pseudorandom numbers successfully pass the DIEHARD tests (a set of statistical tests to determine the qualities of random numbers). The distribution of patients into groups was carried out using a random envelope method. The drug and placebo were outwardly indistinguishable (packaging, label, and content). Investigators, participants, and all study staff were masked to group assignment.

### 2.4. Procedures

All participants who consented to participate attended a screening visit and were tested for eligibility according to eligibility criteria. Testing included physical examination, checks of vital signs, blood tests for infections, tests for drugs and alcohol, pregnancy test (in female participants) etc.

Participants, who passed screening procedures, were accommodated in the hospital on the evening before vaccination. In the morning before vaccination, venous blood samples were collected for subsequent immunogenicity analyzes (day 0 and 21). After vaccination, the participants stayed in the hospital under the supervision of medical personnel over the next two days.

The vaccine GamEvac-Combi is based on two recombinant viral vectors: rVSV-GP and rAd5-GP, both expressing Zaire Ebolavirus glycoprotein. Full doses (1 ml/dose) are 2.5 × 10^7^ plaque forming units of recombinant vesicular stomatitis virus (component A) and 2.5 × 10^11^ viral particles of recombinant human adenovirus 5 serotype (component B). The placebo consists of the vaccine buffer compositions (buffers A and B, accordingly) but without the recombinant viral vectors, made up to equal the vaccine volume. Both vaccine components and placebo were developed, manufactured, and stored by Gamaleya National Research Center of Epidemiology and Microbiology (Moscow, Russia) according to GMP standards.

The vaccine (rVSV-GP at day 0, rAd5-GP at day 21) or placebo were administered intramuscularly into the deltoid muscle with a 21-day interval between doses. An outpatient visit took place on the day 7 after the administration of rVSV-GP. Subsequent visits were planned on 28 day, 42 day, 3 months, 6 months and 12 months. Systemic and local reactions were monitored via examination and tests at visits, through volunteer’s anamnesis and regular diary signs during the whole period of the study.

Blood samples to obtain sera for antibody analysis were collected on days 0, 21, 28, 42; 3, 6 and 12 months. Blood samples for PBMC isolation and subsequent IFN-γ response analyzes were collected at days 0 and

### 2.5. Outcomes

Primary outcome was to determine immunogenicity and safety of GamEVac-Combi in healthy adult participants. Safety was assessed by monitoring of adverse events in vaccinated and placebo groups, Systemic and local post-vaccination reactions in participants were registered throughout the study (up to 12 months after vaccination). Immunogenicity study in vaccinated and placebo groups included evaluation of GP-specific antibody titers at 21, 28, 42 days, 3, 6, 12 months after the vaccination, evaluation of GP-specific T-cell IFN-γ response at day 28. Given the fact that immune response to Plasmodium falciparum antigens may significantly influence the effectiveness of subsequent vaccination, statistical analysis of the subgroup of participants who received antimalarial treatment between the administration of components A and B or after vaccination during follow-up were carried out separately.

### 2.6. Antibody immune response

Sera samples for antibody response were collected in all enrolled participants, who attended corresponding visits on 0, 21, 28, 42 days 3, 6, 12 months. Titers of glycoprotein-specific antibodies in sera samples of participants were evaluated by enzyme-linked immunosorbent assay (ELISA) as described before [3]. Briefly, ELISA plates were coated overnight with a recombinant ZEBOV GP subtype Zaire, strain H.sapiens-wt/GIN/2014/Kissidougou-C15 (Sino Biological, SB40442), washed with phosphate buffer saline containing 0.1% Tween-20 (PBST) and blocked. Diluted sera samples were added in the plates and incubated for 2h, the plates were washed and HRP-conjugated anti-human IgG were added. After the wash, TMB was added, the reaction was stopped by adding H2SO4 and OD was detected at 450 nm (baseline 620 nm). The sample (day 21-360) was evaluated as positive if its optical density was ≥2 times greater than the average optical density of pre-vaccination (day 0) sample of the volunteer within the same dilution. The titer was considered as reciprocal value of maximal dilution which meets the above criteria. Initial dilution of samples was 1:50, negative samples were assigned the titer of 12.5 for statistical analysis. Relative amounts of glycoprotein-specific antibodies are reported as geometric mean end-point titers (GMT) with 95% confidence intervals.

### 2.7. Neutralizing antibody response

Neutralizing antibody titers were determined at days 0 and 42 by plaque pseudoneutralization assay with rVSV-GP. Briefly, Vero E6 cells were seeded in 24-well plates on the day prior to assay to yield monolayer on the day of the assay. Sera samples were heat-inactivated (56oC 30 min) and diluted in DMEM with 2% heat-inactivated fetal bovine serum (Capricorn). Then 100 PFU of rVSV-GP was added to diluted samples (or to equal volume of DMEM for viral dose control), mixed, incubated at 37oC for 1 hour and transferred to the plates with cell monolayer. The plates were incubated for 1 hour (37oC, 5% CO2), then medium was discarded and cells were immediately coated with DMEM-CMC pre-heated to 37oC. After 2 days the plates were washed with sterile isotonic saline solution, fixed and stained with 2.5% Crystal Violet solution (PanReac AppliChem) containing ethanol and formaldehyde (PanReac AppliChem). Neutralization titer was considered as a reciprocal value of maximal dilution of sera sample, which efficiently neutralized at least 50% of PFU, i.e. the mean number of plaques in corresponding wells was ≤2 times lower than the mean number of plaques in viral control wells. Initial dilution of samples was 1:10, negative samples were assigned the titer of 2.5 for statistical analysis.

### 2.8. Cell-mediated immune response

Cell-mediated immune response was assessed in 190 vaccinated and 10 placebo participants at days 0 and 28. Concentration of interferon gamma (IFN-γ) was evaluated by ELISA (Human IFN-gamma Platinum ELISA, BMS228CE, eBioscience) as described before [3]. For this, peripheral blood mononuclear cells (PBMCs) were isolated at day 0 before vaccination and day 28 using Ficoll 1.077 (Paneco) and scattered into 96-well plates. Then recombinant ZEBOV GP subtype Zaire, strain H.sapiens-wt/GIN/2014/Kissidougou-C15 (Sino Biological, 40442-V08B1) was added to stimulate PBMC proliferation. As a positive control, PHA was added to the cells. The culture medium of samples was analyzed after 48 hours incubation to determine IFN-γ concentration. Results are reported as increase in IFN-γ concentration in PBMC samples upon exposure to ZEBOV GP at day 28 compared to day 0 (baseline IFN-γ concentration in GP-stimulated PBMC sample of the participant).

### 2.9. Statistical analysis

The statistical analysis was performed using GraphPad 10 software (v. 10.2.3. GraphPad, USA). The normality of data distribution was analyzed using the Shapiro-Wilks test. Depending on the normality, t-test or the Wilcoxon test was used for analysis of paired values, t-test or the Mann–Whitney test – for unpaired values. Differences were considered significant at p <0.05. Antibody response is reported as geometric mean titers with 95 % confidence intervals (CI), cell-mediated response is reported as median with 95 % confidence intervals. To compare the frequency indicators between groups, the χ^2^ test and, if necessary, Fisher’s exact test were used (if the expected frequency in any of the cells was <5).

## 3. Results

### 3.1. Study Participants

In the Republic of Guinea between August 2017 and December 2018, we recruited and screened 4137 healthy adults for compliance with the criteria, of whom 2000 were randomly assigned to receive vaccine (n=1900) or placebo (n=100; Figure 1). The vaccination was started on August 6, 2017 and completed in December 2018: 1894 participants received first dose of the vaccine and were included in safety analysis, 100 participants received first dose of placebo and were included in safety analysis; 1805 participants were administered both doses of the vaccine (rVSV-GP + rAd5-GP). In Russia 21 healthy adults were screened, 10 volunteers were enrolled and received two doses of the vaccine. Safety set included all participants who received first dose of vaccine/placebo: 1904 vaccinated and 100 placebo participants. Analysis of antibody response included all participants, who attended corresponding visits, IFN-γ response and neutralization analyses included 190 vaccinated participants and 10 participants from placebo group.

**Figure 1.**
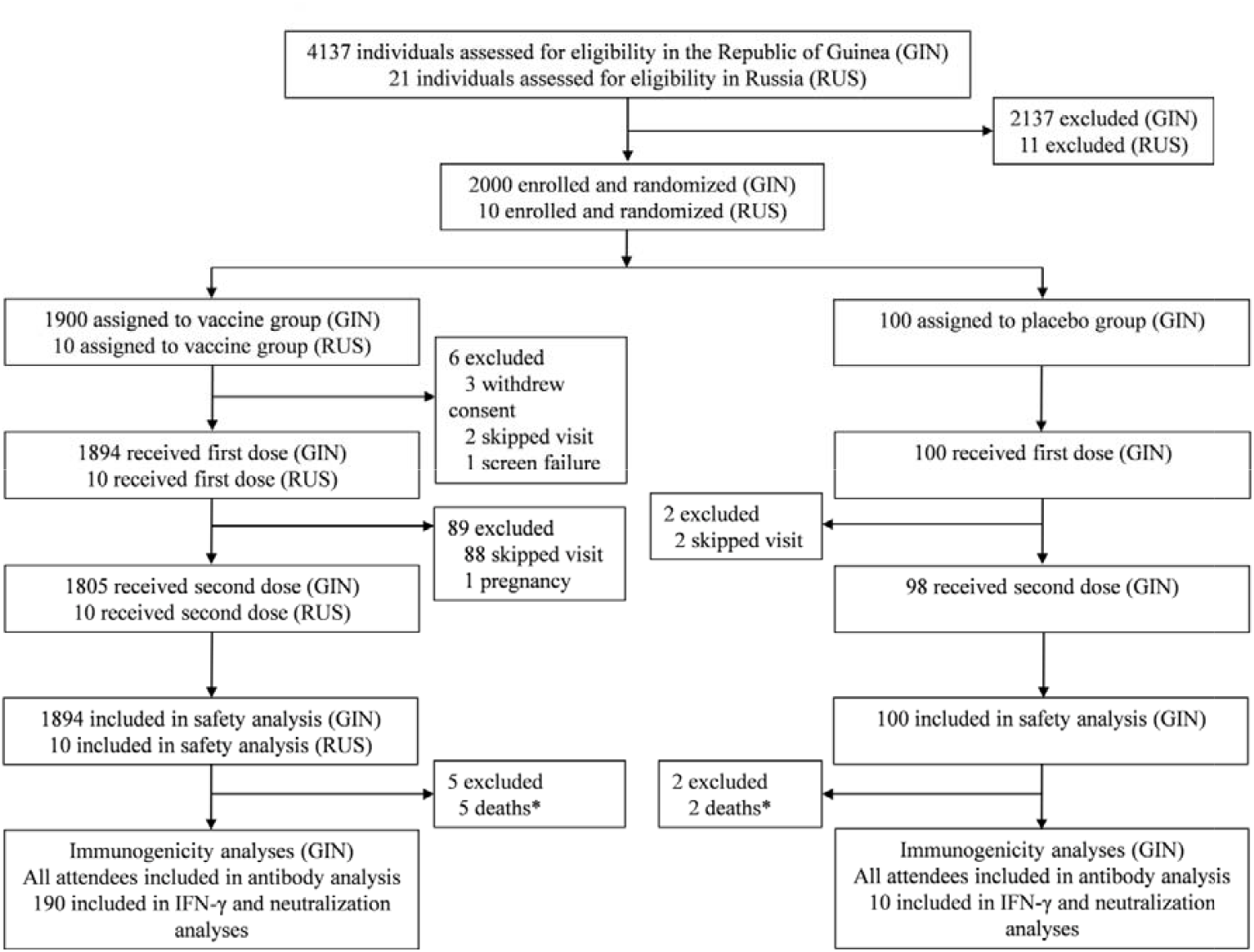
Trial profile. (*) – all reported deaths were not related to received treatment.

Among the participants who received at least one dose, the mean age was 24.5 years (SD 6.8) in the vaccine group and 24.0 years (SD 6.5) in the placebo group; the distribution by sex was similar between the two groups (Table 1).

**Table 1.** Baseline characteristics of participants. Data is indicated n (%) and mean (SD).

|  | **Vaccine (n = 1904)** | **Placebo (n = 100)** |
| --- | --- | --- |
| **Sex** |  |  |
| Male | 1584 (83.2%) | 83 (83%) |
| Female | 315 (16.5%) | 17 (17%) |
| missed data | 5 (0.3%) | - |
| **Age, years** | 24.5 (6.8) | 24.0 (6.5) |
| **Age group, years** |  |  |
| 18-25 | 1366 (71.7%) | 77 (77%) |
| 26-30 | 299 (15.7%) | 13 (13%) |
| 31-35 | 94 (4.9%) | 2 (2%) |
| 36-40 | 52 (2.7%) | 4 (4%) |
| 41-45 | 36 (1.9%) | 2 (2%) |
| 46-50 | 29 (1.5%) | 2 (2%) |
| 51-60 | 20 (1.1%) | 0 (0%) |
| missed data | 8 (0.4%) | - |
| **Body weight, kg** | 63.3 (9.4) | 63.5 (8.5) |
| missed data | 6 (0.3%) | - |
| **Height, cm** | 170 (8.2) | 171 (8.9) |
| missed data | 7 (0.4%) | - |
| **Body-mass index, kg/m<sup>2</sup>** | 22.1 (3.3) | 21.8 (2.9) |
| missed data | 7 (0.4%) | - |

### 3.2. Safety

Clinical monitoring of adverse events was conducted during the entire study up to 12 months after vaccination. Collected data shows that the frequency and nature of adverse events recorded after the vaccine administration corresponds to the available information on the safety of the drug indicated in the Researcher’s Brochure and official instructions for the use of the vaccine.

During the whole period of the trial 2494 adverse events (AE) were reported in 1001 subjects. Of the 2494 AEs, only 1054 AEs in 561 subjects were attributable to vaccine or placebo administration; however, for 294 AEs, the category of causality was marked as unasessable. During the period of day 21 after the second vaccine/placebo administration to final visit, 1342 adverse events were reported in 616 subjects. Of the 1342 AEs, only 389 AEs in 245 subjects were caused by vaccine or placebo administration; however, for 131 AEs, the category of causality was marked by the study physicians as unasessable.

The majority of AEs associated with vaccine/placebo use (AAE) were mild: 96.97% and 96.94% after 1st and 2nd doses, respectively. At the same time, no AAEs with severity grades 3-5 (Severe, Life-threatening or disabling, Death related) were reported (Table A1). Most of the AEs resolved without sequelae. To relieve 60,1% of AEs, it was necessary to use medications or other types of therapy.

In the structure of frequent adverse events, systemic reactions are represented by: an increase in body temperature between 37.2°C and 38.0°C, fever, headache, muscle pain, joint pain, chills, weakness; local reactions – pain and hyperemia at the injection site (Table 2). During the whole period of observation we registered 194 cases of malaria in 172 participants: 183 cases in 162 vaccinated participants and 11 cases in 10 participants in placebo group. No significant difference of frequency of occurrence of adverse events in participants with and without malaria treatment was reported. There were no cases requiring emergency medical care due to administration of GamEvac-Combi vaccine. No deaths or life-threatening or disabling, or severe adverse events, or adverse events resulting in withdrawal from the study related to the vaccine administration were reported (Table A2).

**Table 2.** Frequent adverse events associated with vaccine/placebo administration (AAE). AAEs reported during the whole study are shown in the table as a % of subjects with reported AE.

| Adverse event | Frequency of occurrence (%) |  |
| --- | --- | --- |
| Systemic reactions | Vaccine (n = 1904) | Placebo (n = 100) |
| hyperthermia | 18,49 | 10,00 |
| headache | 11,45 | 10,00 |
| muscle pain | 6,88 | 3,00 |
| joint pain | 2,05 | 3,00 |
| chills | 1,31 | 2,00 |
| weakness | 1,00 | 1,00 |
| Local reactions |  |  |
| pain at the injection site | 2,42 | 2,00 |
| hyperemia at the injection site | 0,16 | 1,00 |

### 3.3. Antibody immune response

To evaluate post-vaccination antibody immune response, we analyzed GP-specific antibody titers by ELISA on 21, 28, 42 days, 3, 6 and 12 months (Figure 2). Sample sets included all participants, who attended corresponding visits: day 21 (n = 1805), day 28 (n = 1775), day 42 (n = 1756), 3 months (n = 1722), 6 months (n = 1686), 12 months (n = 1743). Geometric mean titers (GMT) of GP-specific antibodies and 95% CI were: 21 day – 144.9 (132.6-158.4), 28 day – 9192 (95% CI 8586-9840), 42 day – 9345 (95% CI 8759-9971), 3 months – 3364 (95% CI 3149-3593), 6 months – 1003 (95% CI 941.0-1070), 12 months – 650.0 (95% CI 607.0-696.1). All vaccinated participants showed significant increases in titers at day 28 (7 days after rAd5-GP boost vaccination) compared to day 21 (p < 0.0001). GP-specific antibody levels in vaccinated participants peaked at days 28-42 and gradually decreased over the whole observation period, but stayed significantly higher up to 12 months after vaccination than before booster vaccination (p < 0.0001). Analysis of GP-specific IgG in the group of vaccinated volunteers showed that on day 21 of the study, seroconversion rate (4+ folds increase) was 67.9%, day 28 – 97.7%, day 42 – 97.9%, day 90 – 97.3%, day 180 – 96.3%, day 365 – 94.5%.

**Figure 2.**
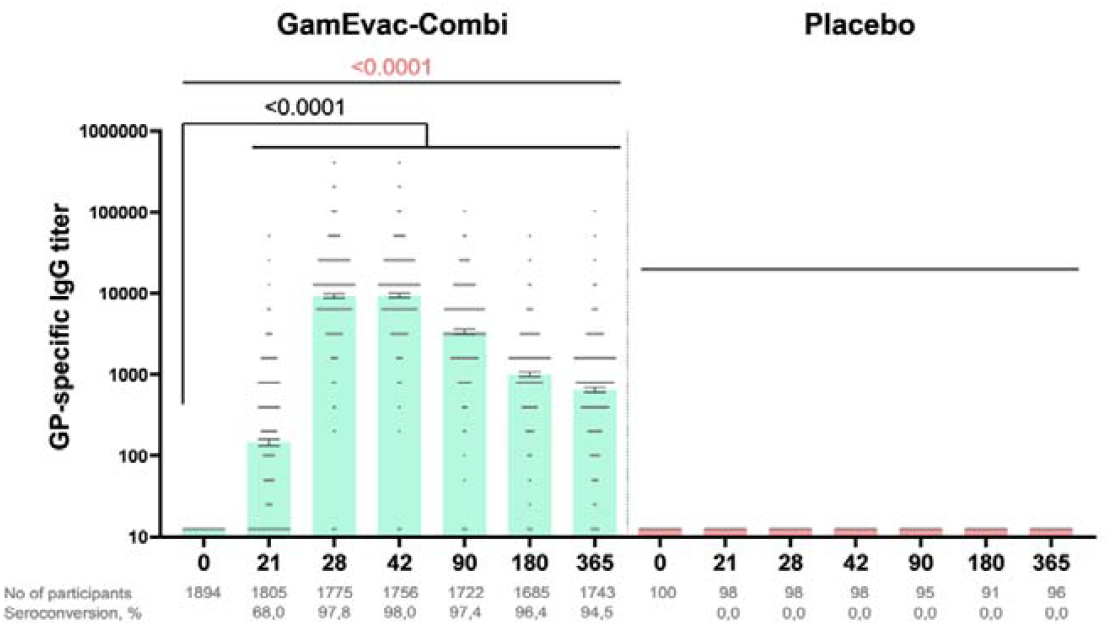
Humoral immune response in vaccinated participants. GP-specific antibody titers at days 0, 21, 28, 42, 90, 180 and 365, as measured by ELISA are shown. Bars show GMT, 95% CI are marked by whiskers. Visits on 3, 6, 12 months are marked as day 90, 180, 65 correspondingly. Differences between IgG titers at days 21 and 28, 42, 90, 180, 365 in comparison with day 0 were calculated with Wilcoxon test (black color). Differences within groups were calculated by ANOVA test (pink color).

We also analyzed antibody response in participants with malaria (since taking antimalarial drugs may affect the formation of a post-vaccination immune response) and no significant difference in antibody responses in participants with and without malaria treatment was reported at days 28-180 post vaccination (Figure B1).

### 3.4. Cell-mediated immune response

Cell-mediated response was evaluated in 190 vaccinated participants and 10 placebo participants by measuring IFN-γ production in PBMC samples by ELISA on days 0 and 28 and reported as increase in IFN-γ concentration in EBOV GP-stimulated PBMCs at day 28 against day 0 in vaccine group (Figure 3). Median IFN-γ concentration in vaccinated volunteers was 0 (interquartile range [IQR] 0–2.0) at day 0 and 26.8 (IQR 4.8– 126.8) at day 28. This indicates formation of GP-specific T-cell response at day 7 after boost vaccination. IFN-γ response was not detected in placebo group.

**Figure 3.**
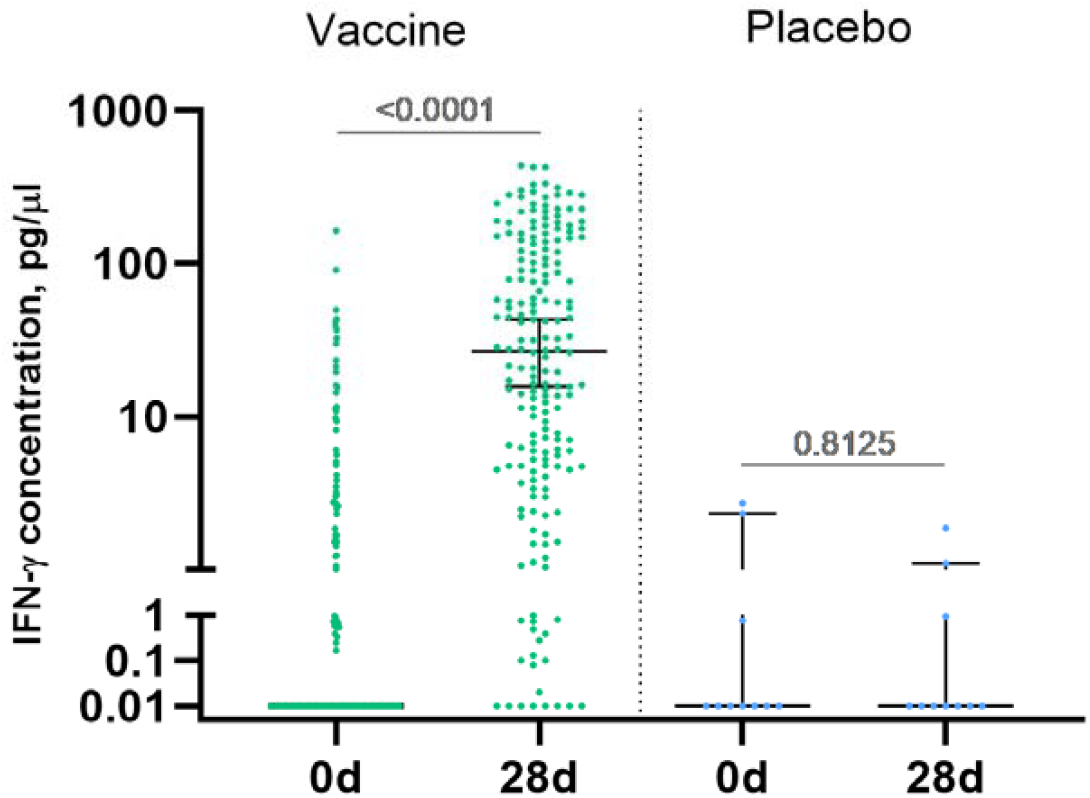
IFN-γ response to Zaire Ebolavirus glycoprotein (GP) in participants. Peripheral blood mononuclear cells were isolated from blood samples of vaccinated participants (n = 190) at days 0 and 28 and stimulated with EBOV-GP. IFN -γ concentration in samples was measured by ELISA. Increase in IFN-γ production (pg/μl) by GP-stimulated PBMCs compared to intact PBMCs at days 0 and 28 is shown on a logarithmic scale. Median is marked with bars, IQR is marked with whiskers. Difference between IFN -γ concentration at days 0 and 28 was calculated with Wilcoxon test.

### 3.5. Neutralizing response

Additionally, neutralizing immune response was evaluated in 190 vaccinated participants and 10 placebo participants. Neutralizing antibody titers were determined in plaque pseudoneutralization assay. Vaccinated participants showed significant neutralizing response at day 42 (p<0,0001) (Figure 4): geometric mean titer of neutralizing antibodies (NtAb) in vaccinated individuals was 32.61, seroconversion rate – 96.3%.

**Figure 4.**
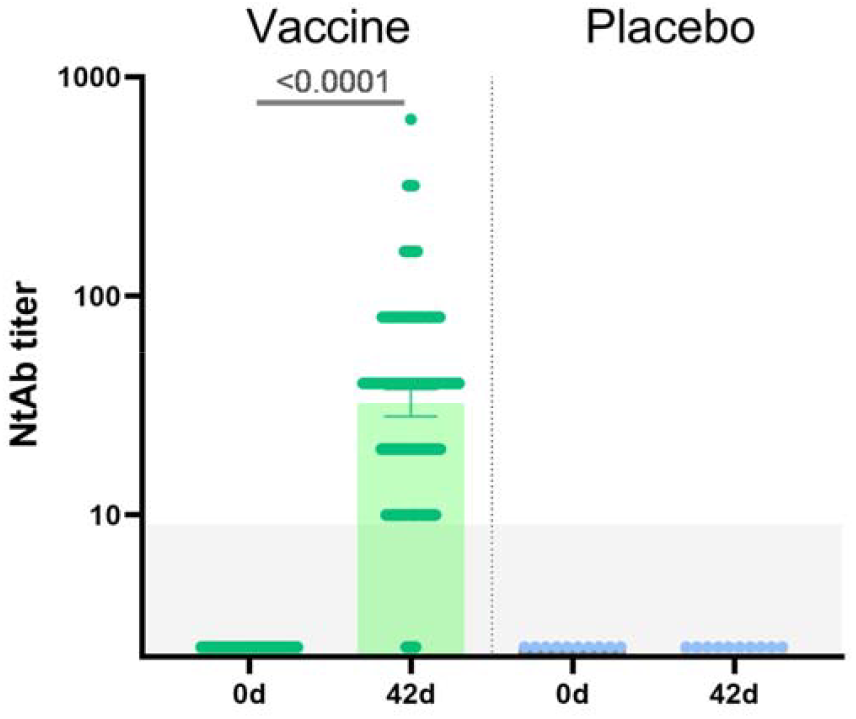
Neutralizing antibodies in sera samples of participants from vaccine group (n = 190) and placebo group (n = 10) at days 0 and 42. Neutralizing antibody (NtAb) titers were determined in plaque pseudoneutralization assay with rVSV-GP. Bars show GMT, 95% CI are marked by whiskers. Difference between NtAb titers at days 0 and 42 was calculated with Wilcoxon test (* p < 0.0001).

## 4. Discussion

In December 2015, GamEvac-Combi – a combined EVD vaccine based on the recombinant viral vectors rVSV and rAd5 - was registered in the Russian Federation after successful completion of phase 1-2 clinical trial [6]. In August 2017 – December 2019 post-registration double-blind randomized placebo-controlled clinical trial of the vaccine was conducted in the Republic of Guinea and in Russia. The primary outcomes of the study were to evaluate safety and immunogenicity of the GamEvac-Combi vaccine in healthy adults. To evaluate vaccine safety, systemic and local reactions were monitored during all study in 1900 vaccinated participants and 100 participants, who received placebo, in the Republic of Guinea and in 10 vaccinated participants in Russia. Antigen-specific humoral immune response was studied in all attended participants from vaccinated and placebo groups up to 12 months after vaccination, IFN-γ responses at day 28 were analyzed in 190 participants from vaccine group and 10 participants from placebo group.

Current study presents the final results of clinical trial of rVSV and rAd5 vaccine against Ebola virus disease. The vaccine showed a favorable safety profile, which is in line with phase 1-2 clinical trial results [6]. No severe, life-threatening or disabling, or death related adverse events, associated with vaccination were reported. Most of the adverse events, associated with the vaccine administration, were mild. All registered adverse events were solicited, occurred within the first days after vaccination and resolved within the next 1-2 days. No vaccine-related deaths were reported. When comparing the immunogenicity data obtained in the present study and in the phase 1-2 study, we noticed that in general, the level of immune response in the population of the Republic of Guinea was higher: for example, on day 42, the geometric mean titer of GP-specific antibodies in phase 1-2 clinical trial was 3277 (95% CI 2401-4473) [6], while in this study the antibody level at day 42 was 9349 (95% CI 8762-9975).

According to heterologous prime-boost approach, component A (rVSV) was administrated at day 0 and component B (rAd5) – at day 21, which resulted in significant boost of antibody levels at day 28, prominent peak of GP-specific antibodies at day 42 and a year-long persistence of antibody response. Vaccination with rVSV and rAd5 also induced significant IFN-γ response 7 days after the boost.

An important advantage of the GamEvac-Combi vaccine is the use of a heterologous prime-boost vaccination approach. The kinetics of the immune response in volunteers is adequate: after vaccination, a peak response is detected at 28-42 days, and then a systematic decrease in the level of antibodies at 6-12 months. A year after vaccination, specific antibodies were detected in the blood serum of more than 94% of volunteers. The seroconversion rate is significantly higher than with a single vaccination with registered vaccines based on recombinant chimpanzee adenovirus (ChAd3-EBO-Z) and recombinant vesicular stomatitis virus (rVSVΔG-ZEBOV-GP): studies in Liberia showed that one year after vaccination % of volunteers with IgG response was 63.5% in the ChAd3-EBO-Z group and 79.5% in the rVSVΔG-ZEBOV-GP group [7]. Thus, vaccination with GamEvac-Combi allows to form a long-term intense immune response.

Our study has some limitations. First, predominantly male participants and mainly of young age. Second, the lack of laboratory correlates of protection makes it difficult to interpret the clinical significance of changes in antibody levels. Thirdly, within the framework of this study, it was not possible to assess the epidemiological effectiveness of the vaccine, since no cases of EVD were recorded in the region during the study.

Heterologous prime-boost approach was used in development of Sputnik V (Gam-COVID-Vac) vaccine against COVD-19. Gam-COVID-Vac consists of two doses: rAd26-based vector is used for the prime and rAd5-based vector is used for the boost 21 days later. This approach has proven to effectively induce antigen-specific immune response in multiple clinical studies [8, 9]. GamEvac-Combi uses the same approach with rVSV for priming and rAd5 for boosting immune response. Recombinant replication-competent VSV induces rapid innate immunity activation, which was shown to contribute to early protection against lethal EBOV infection in primate studies [10]. The vaccine rVSV-ZEBOV has shown high efficacy in clinical studies and to date is the only vaccine with reported efficacy in EVD outbreaks [11, 12]. The findings from this trial might contribute to data on vaccine-induced immune responses against Ebola virus disease.

## Data Availability

All data produced in the present study are available upon reasonable request to the authors

## Supplementary Materials

The following supporting information can be downloaded at: www.mdpi.com/xxx/s1.

## Author Contributions

Conceptualization, Denis Logunov, Sergei Borisevich, Boris Naroditsky and Alexander Gintsburg; Data curation, Georgiy Prokhorov, Alexander Khovaev, Tatiana Demidova, Nikolai Malishev, Liliya Merkulova, Olga Voronina, Irina Fedyakina, Lidiya Kisteneva, Lyudmila Kolobukhina, Dmitry Mishin, Aleksandr Elakov, Ekaterina Ermolova, Kirill Krasnoslobodtsev, Viktor Larichev, Irina Kruzhkova, Egor Burmistrov, Anna Sheremet, Elizaveta Tokarskaya, Roman Gudkov, Oleg Maltsev, Ilnur Gabdrahmanov, Anton Barsukov, Vladislav Vashchenkov, Nikolaj Demianenko, Sergej Ignatev, Konstantin Asyamov, Nikolay Kirichenko, Andrej Lyubimov, Igor Volkov, Evgenij Kryukov, Nikolay Bazarnov and Victoria Kolodyazhnaya; Formal analysis, Inna Dolzhikova, Anna Kovyrshina, Alina Dzharullaeva, Amir Tukhvatulin, Maria Ordzhonikidze, Dmitrii Shcheblyakov, Ilias Esmagambetov, Maksim Shmarov and Alexander Gromov; Funding acquisition, Alexander Gintsburg; Investigation, Inna Dolzhikova, Anna Kovyrshina, Alina Dzharullaeva, Alina Erokhova, Daria Grousova, Amir Tukhvatulin, Fatima Izhaeva, Olga Zubkova, Dmitrii Shcheblyakov, Ilias Esmagambetov, Maksim Shmarov, Natalia Tukhvatulina, Nikolaevich Shcherbinin, Irina Tutykhina, Dmitrii Reshetnikov, Svetlana Syromyatnikova, Dmitry Chifanov, Alexander Andrus and Dmitry Kutaev; Methodology, Denis Logunov, Inna Dolzhikova, Alina Dzharullaeva, Amir Tukhvatulin, Yana Simakova, Olga Zubkova and Dmitrii Shcheblyakov; Project administration, Denis Logunov and Yana Simakova; Resources, Mamadou Boiro, Nadezhda Lubenec, Alexander Semikhin, Aleksandr Fisun, Bogdan Kotiv, Dmitrij Ovchinnikov, Evgenij Ivchenko, Konstantin Zhdanov, Sergej Zakharenko, Aleksandr Solovev, Andrej Ivanov, Vitalij Sukachev, Evgenij Kryukov and Elena Kolomoets; Supervision, Sergei Borisevich and Boris Naroditsky; Visualization, Inna Dolzhikova, Anna Kovyrshina and Maria Ordzhonikidze; Writing – original draft, Inna Dolzhikova and Anna Kovyrshina; Writing – review & editing, Inna Dolzhikova, Amir Tukhvatulin and Dmitrii Shcheblyakov. All authors will be informed about each step of manuscript processing including submission, revision, revision reminder, etc. via emails from our system or assigned Assistant Editor.

## Institutional Review Board Statement

The study was conducted in accordance with the Declaration of Helsinki, and approved by the appropriate national and local competent authorities, including the Ethics Committee of the Ministry of Health of the Russian Federation (#119 16/02/2016, additional #155 19/09/2017, #203 19/11/2019) and Ethics Committee of the Ministry of Health of Republic of Guinea (76/CNERS/16 20/06/2016, № 064/CNERS/17 07/05/2017, № 041/CNERS/18 09/03/2018).

## Informed Consent Statement

Informed consent was obtained from all subjects involved in the study.

## Data Availability Statement

Anonymous participant data will be available upon request to the corresponding author. Proposals will be reviewed and approved by the sponsor, security department, researcher, and staff on the basis of scientific merit and absence of competing interests. Once the proposal has been approved, data can be transferred through a secure online platform after the signing of a data access agreement and a confidentiality agreement.

## Acknowledgments

We thank the people in Kindia for their participation, the entire staff of Medical service CBK RUSAL, Research Center for Epidemiology, Microbiology and Medical Care.

## Supplementary material

### Supplementary Authors List – GamEvac-Combi trial group

|  |  |
| --- | --- |
| Konomou Kpakile Victor | Medical service CBK RUSAL, Research Center for Epidemiology, Microbiology and Medical Care, B.P:146, Kindia Republic of Guinea, CREMS (Pastori) |
| Camara Sekou Bakary |  |
| Diallo Mamadou Yero |  |
| Kolie Moussa Claver |  |
| Camara Ibrahima Aissatou |  |
| Soumah Idrissa |  |
| Gbamou N'yereke Jerome |  |
| Shishina Anna Valentinovna | Federal state budgetary military educational institution of higher education «Military Medical Academy named after S.M.Kirov» of the Ministry of defence of the Russian Federation |
| Beliaevskaia Irina Vladimirovna |  |
| Komissarova Galina Mikhajlovna |  |
| Yaroczkaya Olga Aleksandrovna |  |
| Glazkova Marina Anatolyevna | National Research Centre of Epidemiology and Microbiology named after Honorary Academician N. F. Gamaleya, Ministry of Health of Russian Federation |
| Telegina Anastasia Aleksandrovna |  |
| Pushkarev Andrey Yurevich |  |

**Table A1.** Systemic and local adverse events by grade, associated with vaccine/placebo administration (AAE). AAEs reported during the whole study are shown in the table as a number of subjects with reported AE and % of the group.

|  | Grade | Vaccine (n = 1904) |  | Placebo (n = 100) |  | Overall (n = 2004) |  | p-value |
| --- | --- | --- | --- | --- | --- | --- | --- | --- |
| Overall AEs | Mild | 532 | 27,94% | 18 | 18,00% | 550 | 27,45% | 0,0299 |
|  | Moderate | 21 | 1,10% | 0 | 0,00% | 21 | 1,05% | 0,2911 |
| Systemic and local reactions | **Mild** | **395** | **20,75%** | **13** | **13,00%** | **408** | **20,36%** | **0,0608** |
|  | **Moderate** | **14** | **0,74%** | **0** | **0,00%** | **14** | **0,70%** | **0,3895** |
| Hyperthermia | Mild | 342 | 17,96% | 10 | 10,00% | 352 | 17,56% | 0,0414 |
|  | Moderate | 13 | 0,68% | 0 | 0,00% | 13 | 0,65% | 0,4071 |
| Pain (local) | Mild | 45 | 2,36% | 2 | 2,00% | 47 | 2,35% | 0,8149 |
|  | Moderate | 1 | 0,05% | 0 | 0,00% | 1 | 0,05% | 0,8187 |
| Chills | Mild | 24 | 1,26% | 2 | 2,00% | 26 | 1,30% | 0,5241 |
|  | Moderate | 1 | 0,05% | 0 | 0,00% | 1 | 0,05% | 0,8187 |
| Asthenia | Mild | 19 | 1,00% | 1 | 1,00% | 20 | 1,00% | 0,9984 |
| Malaise | Mild | 6 | 0,32% | 0 | 0,00% | 6 | 0,30% | 0,5740 |
| Erythema (local) | Mild | 2 | 0,11% | 1 | 1,00% | 3 | 0,15% | 0,0241 |
|  | Moderate | 1 | 0,05% | 0 | 0,00% | 1 | 0,05% | 0,8187 |
| Feeling of fever (subjective) | Mild | 1 | 0,05% | 0 | 0,00% | 1 | 0,05% | 0,8187 |
| Fatigue | Mild | 1 | 0,05% | 0 | 0,00% | 1 | 0,05% | 0,8187 |
| Nervous system | **Mild** | **216** | **11,34%** | **10** | **10,00%** | **226** | **11,28%** | **0,6786** |
|  | **Moderate** | **3** | **0,16%** | **0** | **0,00%** | **3** | **0,15%** | **0,6912** |
| Headache | Mild | 215 | 11,29% | 10 | 10,00% | 225 | 11,23% | 0,6900 |
|  | Moderate | 3 | 0,16% | 0 | 0,00% | 3 | 0,15% | 0,6912 |
| Dizziness | Mild | 17 | 0,89% | 0 | 0,00% | 17 | 0,85% | 0,3426 |
| Muscle, skeletal and connective tissue | **Mild** | **156** | **8,19%** | **4** | **4,00%** | **160** | **7,98%** | **0,1316** |
|  | **Moderate** | **1** | **0,05%** | **0** | **0,00%** | **1** | **0,05%** | **0,8187** |
| Myalgia | Mild | 130 | 6,83% | 3 | 3,00% | 133 | 6,64% | 0,1339 |
|  | Moderate | 1 | 0,05% | 0 | 0,00% | 1 | 0,05% | 0,8187 |
| Arthralgia | Mild | 39 | 2,05% | 3 | 3,00% | 42 | 2,10% | 0,5173 |
| Bone pain | Mild | 2 | 0,11% | 0 | 0,00% | 2 | 0,10% | 0,7457 |
| Back pain | Mild | 1 | 0,05% | 0 | 0,00% | 1 | 0,05% | 0,8187 |
| Musculoskeletal discomfort | Mild | 1 | 0,05% | 0 | 0,00% | 1 | 0,05% | 0,8187 |
| Local (muscle) pain | Mild | 1 | 0,05% | 0 | 0,00% | 1 | 0,05% | 0,8187 |
| Skin and subcutaneous tissues | **Mild** | **21** | **1,10%** | **0** | **0,00%** | **21** | **1,05%** | **0,2911** |
|  | **Moderate** | **2** | **0,11%** | **0** | **0,00%** | **2** | **0,10%** | **0,7457** |
| Itch | Mild | 16 | 0,84% | 0 | 0,00% | 16 | 0,80% | 0,3574 |
| Rash | Mild | 4 | 0,21% | 0 | 0,00% | 4 | 0,20% | 0,6464 |
|  | Moderate | 2 | 0,11% | 0 | 0,00% | 2 | 0,10% | 0,7457 |
| Hyperhidrosis | Mild | 2 | 0,11% | 0 | 0,00% | 2 | 0,10% | 0,7457 |
| **Gastrointestinal** | **Mild** | **11** | **0,58%** | **0** | **0,00%** | **11** | **0,55%** | **0,4460** |
| Nausea | Mild | 7 | 0,37% | 0 | 0,00% | 7 | 0,35% | 0,5436 |
| Vomit | Mild | 2 | 0,11% | 0 | 0,00% | 2 | 0,10% | 0,7457 |
| Abdominal pain | Mild | 2 | 0,11% | 0 | 0,00% | 2 | 0,10% | 0,7457 |
| Dyspepsia | Mild | 1 | 0,05% | 0 | 0,00% | 1 | 0,05% | 0,8187 |
| **Infections and infestations** | Mild | 8 | 0,42% | 0 | 0,00% | 8 | 0,40% | 0,5160 |
|  | Moderate | 1 | 0,05% | 0 | 0,00% | 1 | 0,05% | 0,8187 |
| Malaria | Mild | 2 | 0,11% | 0 | 0,00% | 2 | 0,11% | 0,7457 |
|  | Moderate | 1 | 0,05% | 0 | 0,00% | 1 | 0,05% | 0,8187 |
| Gastroenteritis | Mild | 2 | 0,11% | 0 | 0,00% | 2 | 0,10% | 0,7457 |
| Periodontitis | Mild | 1 | 0,05% | 0 | 0,00% | 1 | 0,05% | 0,8187 |
| Pharyngitis | Mild | 1 | 0,05% | 0 | 0,00% | 1 | 0,05% | 0,8187 |
| Typhoid fever | Mild | 1 | 0,05% | 0 | 0,00% | 1 | 0,05% | 0,8187 |
| Nasopharyngitis | Mild | 1 | 0,05% | 0 | 0,00% | 1 | 0,05% | 0,8187 |
| **Cardiac** | **Mild** | **4** | **0,21%** | **0** | **0,00%** | **4** | **0,20%** | **0,6464** |
|  | **Moderate** | **3** | **0,16%** | **0** | **0,00%** | **3** | **0,15%** | **0,6912** |
| Sinus tachycardia | Mild | 1 | 0,05% | 0 | 0,00% | 1 | 0,05% | 0,8187 |
|  | Moderate | 2 | 0,11% | 0 | 0,00% | 2 | 0,10% | 0,7457 |
| Bradycardia | Mild | 1 | 0,05% | 0 | 0,00% | 1 | 0,05% | 0,8187 |
|  | Moderate | 1 | 0,05% | 0 | 0,00% | 1 | 0,05% | 0,8187 |
| Tachycardia | Mild | 1 | 0,05% | 0 | 0,00% | 1 | 0,05% | 0,8187 |
| Heartbeat (subjective palpitation) | Mild | 1 | 0,05% | 0 | 0,00% | 1 | 0,05% | 0,8187 |
| **Metabolic and nutritional** | **Mild** | **5** | **0,26%** | **0** | **0,00%** | **5** | **0,25%** | **0,6079** |
| Loss of appetite | Mild | 4 | 0,21% | 0 | 0,00% | 4 | 0,20% | 0,6464 |
| Appetite disturbance | Mild | 1 | 0,05% | 0 | 0,00% | 1 | 0,05% | 0,8187 |
| **Changes in laboratory variables** | **Mild** | **3** | **0,16%** | **0** | **0,00%** | **3** | **0,15%** | **0,6912** |
|  | **Moderate** | **1** | **0,05%** | **0** | **0,00%** | **1** | **0,05%** | **0,8187** |
| Increased blood pressure | Mild | 3 | 0,16% | 0 | 0,00% | 3 | 0,15% | 0,6912 |
|  | Moderate | 1 | 0,05% | 0 | 0,00% | 1 | 0,05% | 0,8187 |
| **Mental disorders** | **Mild** | **1** | **0,05%** | **0** | **0,00%** | **1** | **0,05%** | **0,8187** |
| Sleep disorder | Mild | 1 | 0,05% | 0 | 0,00% | 1 | 0,05% | 0,8187 |
| **Respiratory** | **Mild** | **1** | **0,05%** | **0** | **0,00%** | **1** | **0,05%** | **0,8187** |
| Rhinorrhea | Mild | 1 | 0,05% | 0 | 0,00% | 1 | 0,05% | 0,8187 |

**Table A2.** Death related severe adverse events.

| № | Group | Date of Component B administration | Date of SAE manifestation | Death-related records | Death cause | Causality assessment |
| --- | --- | --- | --- | --- | --- | --- |
| 1 | Vaccine | 10.09.2017 | 19.08.2018 | The volunteer informed the medical staff about experienced severe abdominal pain and was sent for consultation with a surgeon at the regional hospital in Kindia named after Alpha Dialo. Data on fatal outcome was received from the surgeon. | peritonitis | unrelated |
| 2 | Placebo | 11.07.2018 | 17.12.2018 | The volunteer did not attend visit 7 (6 months after first administration). According to contacts (friends and relatives), the volunteer died three weeks ago and did not visit medical institutions | unknown | unrelated |
| 3 | Vaccine | 12.08.2018 | May 2019 | According to contacts (relatives), the volunteer committed suicide in May 2019, exact date is unknown. | suicide | unrelated |
| 4 | Vaccine | 29.06.2018 | Apr 2019 | According to contacts (relatives), the volunteer died in April 2019, after fainted. | unknown | unrelated |
| 5 | Placebo | 16.08.2018 | 22.01.2019 | The volunteer did not attend visit 7 (6 months after first administration). According to contacts (relatives), the volunteer died in June 2019, the death cause was not disclosed to the staff. | unknown | unrelated |
| 6 | Vaccine | 01.09.2018 | 27.05.2019 | The volunteer died due to road accident. | road accident | unrelated |
| 7 | Vaccine | 04.08.2018 | 28.05.2019 | The volunteer did not attend visit 7 (6 months after first administration). Cause of the death remains not clear. | unknown | unrelated |

**Figure B1.**
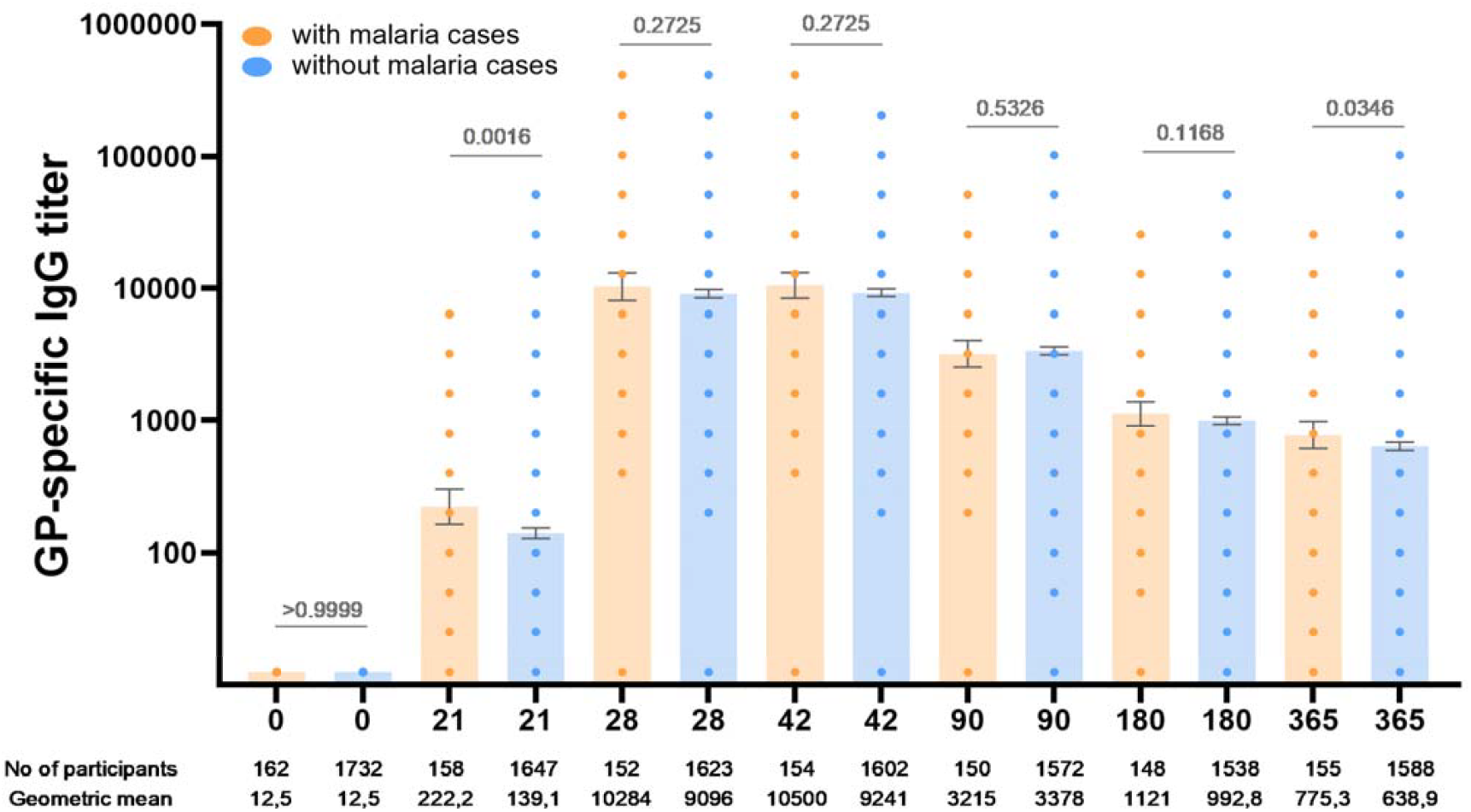
Humoral immune response in vaccinated participants with and without malaria cases. Humoral immune response in vaccinated participants with and without malaria cases. GP-specific antibody titers at days 0, 21, 28, 42, 90, 180 and 365, as measured by ELISA are shown. Bars show GMT, 95% CI are marked by whiskers. Visits on 3, 6, 12 months are marked as day 90, 180, 365 correspondingly. Differences between IgG titers between groups (convalescents and non-convalescent) were calculated with Mann-Whitney test (p value is marked above grey bars).

